# Economic Evaluation of Hypertension screening in Iran using Markov model

**DOI:** 10.1101/2024.04.24.24306273

**Authors:** Rajabali Daroudi, Ali Akbari Sari, Mahmoud Zamandi, Elham Yousefi

## Abstract

**Introduction and purpose:** Hypertension is one of the most common non-communicable diseases in the world. However, in LMCs, there is not enough evidence-based information about the cost-effectiveness of preventive interventions for hypertension. Therefore, the purpose of this study is to conduct an economic evaluation of high blood pressure screening strategies in Iran in 2020.

**Method:** We did an economic evaluation of 9 blood pressure screening strategies, including screening annually or every two or three years from the ages of 30, 40, or 50, using the Markov model. The Markov model was designed and implemented based on the natural history of cardiovascular disease in the 2020 TreeAge Pro software. The quality-adjusted life years and the average cost of high blood pressure screening and treatment per person were estimated from society’s perspective for the lifetime. Input data of the model were derived from published literature, expert opinion, and available data sources

**Findings:** All screening interventions were more costly and more effective compared to no screening. Five strategies, including screening every three years from the age of 50, 40, and 30 years and screening every two years and annually from the age of 30, were undominated. Incremental cost-effectiveness ratios for these strategies ranged from $90.5 to $38,289.57. Probabilistic sensitivity analysis indicated that, at a cost-effectiveness threshold close to one times the GDP per capita, screening every two or three years from age 30 had the highest cost-effectiveness, with probabilities of 0.589 and 0.361, respectively.

**Conclusion:** Based on the findings of the economic evaluation, all screening strategies are more cost-effective compared to no screening, and among the screening strategies, considering about one times the GDP per capita as the cost-effectiveness threshold, a screening strategy every two years, starting at the age of 30, is the most cost-effective strategy.

## Introduction

High blood pressure is one of the most common non-communicable diseases worldwide and is the main risk factor for cardiovascular diseases such as coronary artery disease, stroke, heart failure, arrhythmia, and cardiomyopathy(1).According to studies conducted in 2010, high blood pressure was the cause of 49% of coronary heart disease cases and 62% of stroke cases(2, 3). The prevalence of high blood pressure is increasing in low and middle-income countries including Iran (4, 5).

Studies conducted in different provinces of the Islamic Republic of Iran have shown a wide variety of high blood pressure prevalence (6–8). In Iran, in 2011, about 25.6% and 39.8% of adults aged 25-70 had high blood pressure and pre-hypertension, respectively(9). Although considerable knowledge is available about the epidemiology, pharmacotherapy, and genetics of hypertension, if health systems can not more effectively identify individuals with hypertension and address barriers to healthcare delivery, They are underdiagnosed and not treated(10).

In Iran in 2020, 52.84% of men and 68.02% of women were aware of their high blood pressure (61.48% in both sexes), and among them, only 43.15% of men and 58.73% of women with high blood pressure were treated and received therapeutic drugs (52.02 % in both sexes)(11). Also, in Iran, based on the results of the Global Burden of Diseases study, 6.5 million people are unaware of their high blood pressure disease, half of the people who are aware of it are being treated, and 26% of the people being treated have their blood pressure controlled. By controlling blood pressure, heart attacks and strokes in Iran will be reduced by 25%(12) (13). Preventive interventions such as high blood pressure screening is one of the important interventions to identify people at risk and prevent high blood pressure and minimize the number of affected people and costs. Currently population-based screening for high blood pressure is not implemented in Iran. However, opportunistic screening is done in health centers,.Also, in 2019, a national high blood pressure control campaign was carried out, which, of course, mostly aimed to raise awareness among the society.

Economic evaluation is a structured method that aims to identify, measure, value and compare the costs and consequences of several alternative programs or interventions. and help health policy makers implement more effective health interventions. Economic evaluations help to improve the efficiency of the health system by prioritizing, rationing and optimal allocation of resources and increasing its effectiveness by improving access and equality (14).

Previous studies counducted in low and middle income countries proved the cost-effectiveness of hypertension screening interventions(15) (16–19). So far, the costs and benefits of various high blood pressure screening interventions have not been investigated in Iran.Therefore, the aim of this study is the economic evaluation of different types of blood pressure screening strategies in Iran.

## Methods

This study aimed to assess the cost-effectiveness of different screening strategies for high-blood pressure using a Markov modeling approach in Iran. The Markov model simulated the progression of individuals through health states over time, allowing us to estimate the long-term costs and health outcomes associated with each screening strategy, as well as no screening strategy. The time horizon of the model was life time to capture the long-term costs and health outcomes associated with each screening strategy. The study population consisted of individuals aged 30 years or older. We adopted a societal perspective, considering direct medical and non-medical costs associated with screening, diagnosis, and treatment of high-blood pressure and its complications. Future costs and health outcomes were discounted at an annual rate of 5%.

To compile the data for this study, we utilized results from prior studies, national and international databases, outcomes from the national high blood pressure control campaign, and calculations based on these results, as well as data from Iran’s health insurance. All the data used in this research are publicly available, except for Iran’s health insurance data. Therefore, health insurance data was requested during administrative procedures through the Tehran University of Medical Sciences. The items requested by the researchers were prepared by experts from Iran’s health insurance organization. Throughout the year 2021, on several occasions, each part of the data that was prepared was anonymously made available to the researchers.

### Model structure

We developed a Markov model to simulate the natural history of high-blood pressure and its associated cardiovascular disease (CVD) outcomes over time. The model comprised ten mutually exclusive health states representing different stages of hypertension and CVD events including:

#### Healthy

Individuals in this state have not been diagnosed with hypertension or experienced any CVD events(including CHD or Stroke).

#### Hypertension off treatment

Individuals diagnosed with hypertension but not receiving treatment.

#### Hypertension on treatment

Individuals diagnosed with hypertension and receiving antihypertensive treatment.

#### Post Stroke

Individuals who have experienced a stroke event.

#### Post Myocardial Infarction

Individuals who have experienced a myocardial infarction (heart attack).

#### Post Unstable Angina

Individuals who have experienced unstable angina.

#### Post Stable Angina

Individuals who have experienced stable angina.

#### Post Transient Ischemic Attack

Individuals who have experienced a transient ischemic attack (TIA or “mini-stroke”).

#### CVD Death

Terminal health state representing death related to CVD.

#### Non-CVD Death

Terminal health state representing death from any cause unrelated to CVD.

Each cycle of the model represented a discrete time period during which individuals could transition between health states based on predefined transition probabilities. At the beginning of the model simulation, all individuals were placed in three conditions, including healthy, Hypertension off treatment, and Hypertension on treatment. At the end of each yearly cycle, individuals in each health state may remain in their current state or suffer non-cardiovascular death. Forthermore, those in a healthy state may transition to one of the Hypertension states or encounter a coronary heart disease (CHD) or stroke event. Those in the Hypertension off treatment state, may receive a diagnosis of hypertension and commence antihypertensive treatment, or experience a CHD or stroke event. Similarly, individuals in the hypertension on-treatment state may also experience a CHD or stroke event. In the event of a CHD or stroke, individuals may either perish immediately or transition to one of the post-CVD event states. Individuals within post-CVD event states may continue in that state, undergo another CVD event, or face mortality (Figure 1).

**Figure 1:**
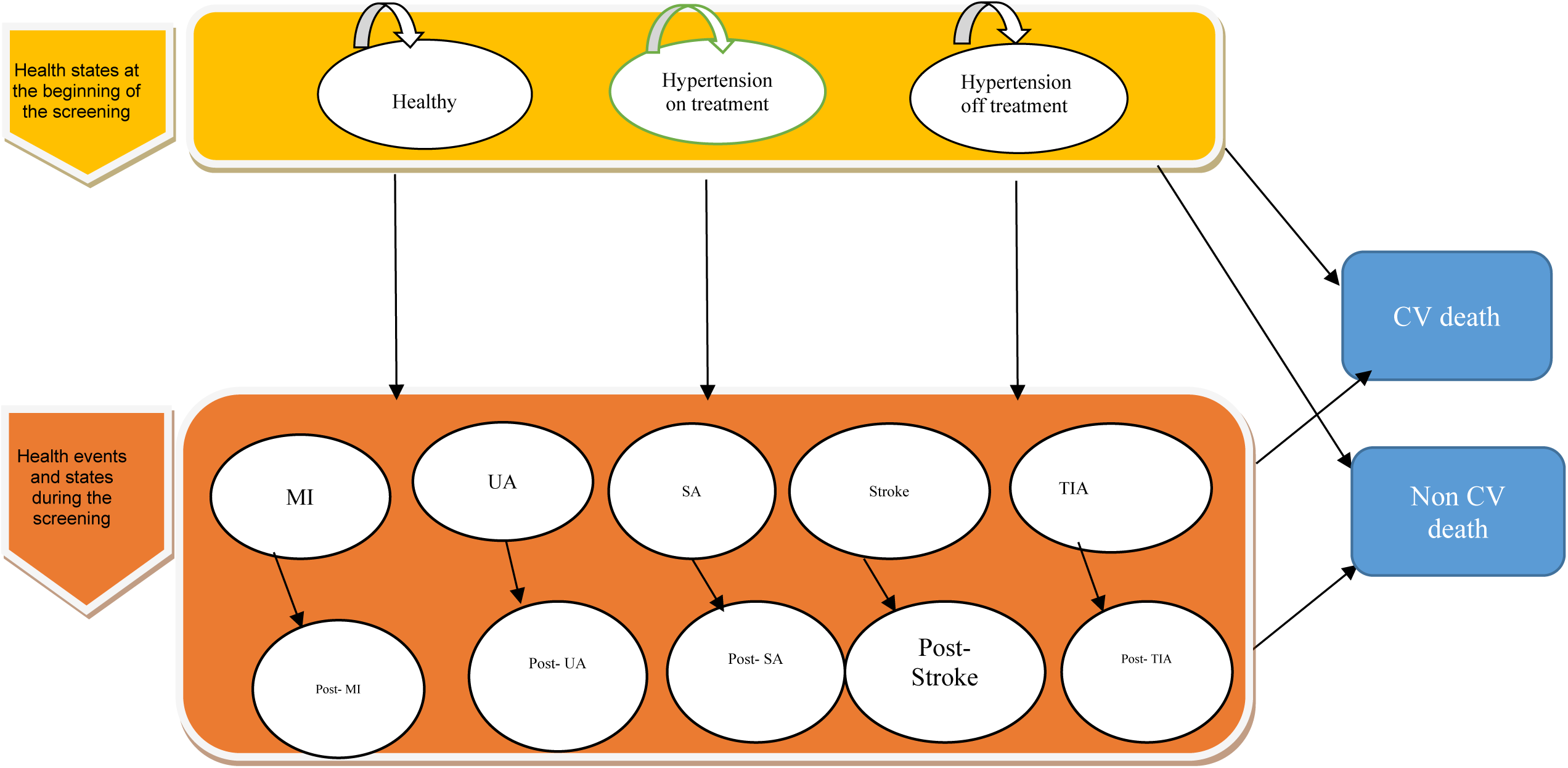
Markov model structure. MI, myocardial infarction; SA, stable angina; TIA, transient ischemic attack; UA, unstable angina.

By conducting screening, people with or at risk of hypertension are diagnosed and treated, so by conducting intervention and screening, people with undiagnosed hypertension are transferred to diagnosed and treated states and As a result, the number of CVD events and death will decreases.

#### Model Assumptions

- Transitions between health states were governed by transition probabilities derived from epidemiological studies, clinical trials, and meta-analyses.
- Individuals could experience multiple CVD events over the course of the simulation.
- The model assumed that individuals remained in the same health state until transitioning to another state or experiencing death.

The Markov model was designed and implemented in the 2020 TreeAge Pro software.

#### Screenign strategies

We evaluated several screening strategies, including:

- No screening : Participants receive no systematic screening for high-blood pressure.
- Annual screening from age 30 to 70 years
- Annual screening from age 40 to 70 years
- Annual screening from age 50 to 70 years
- Biennial Screening from age 30 to 70 years
- Biennial Screening from age 40 to 70 years
- Biennial Screening from age 50 to 70 years
- Three-yearly screening from age 30 to 70 years
- Three-yearly screening from age 40 to 70 years
- Three-yearly screening from age 50 to 70 years

### Model parameters

#### Hypertension Incidence and Prevalence

Epidemiological data concerning the incidence and prevalence of hypertension in Iran, disaggregated by age and sex, were sourced from the STEPs 2016 survey report(20). Additionally, figures representing the proportion of individuals with hypertension who are aware of their condition and the percentage undergoing treatment, categorized by age, were obtained from STEPs 2016 survey report and prior studies conducted in Iran(21).

#### Cardiovascular Risks and Probabilities

In this study, the probability of incidence of cardiovascular diseases in healthy people, individuals with hypertension, and those undergoing hypertension treatment was calculated separately (Table 1) using the relative risks and the incidence rates of each group(22).

**Table 1.**
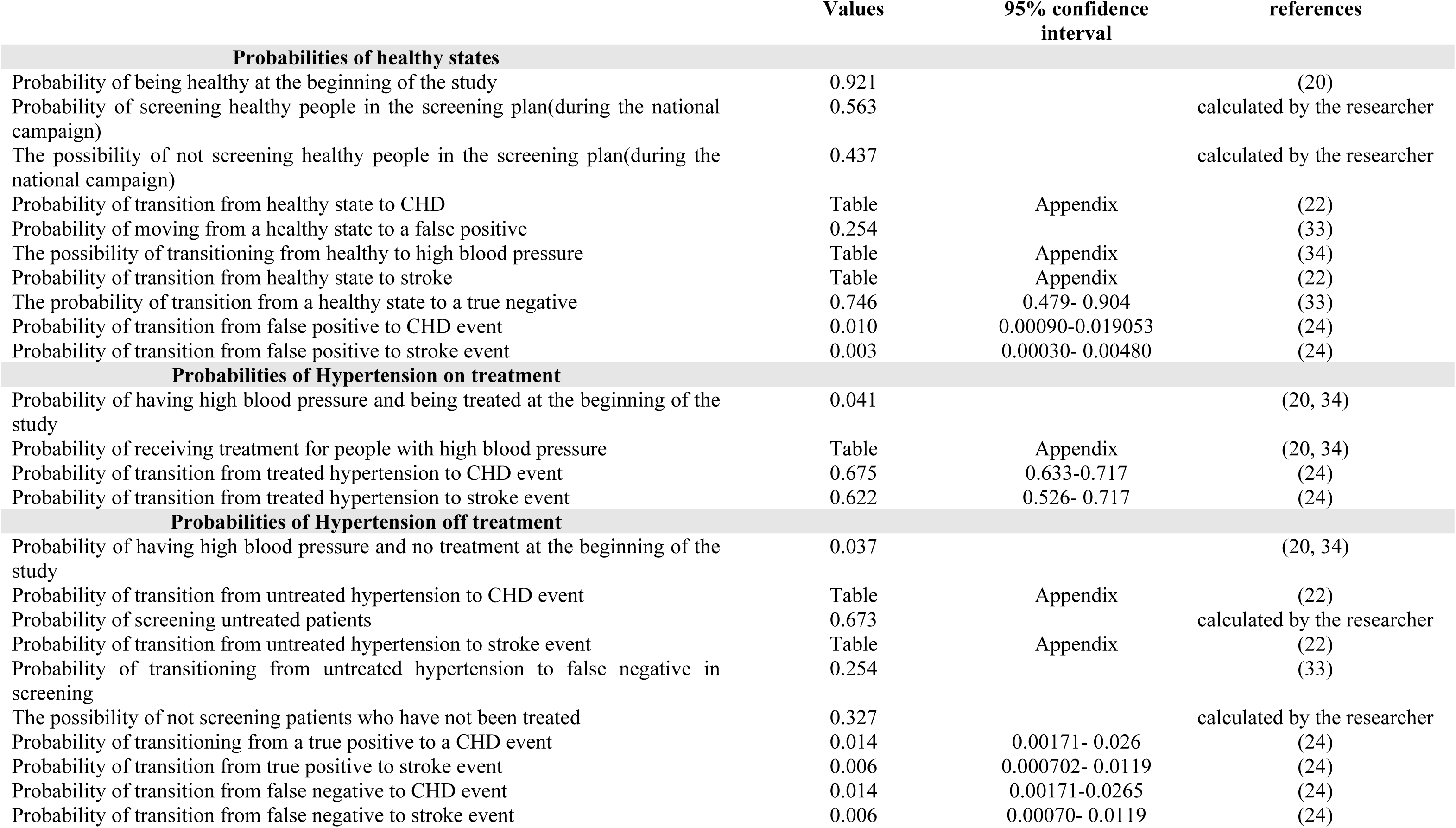
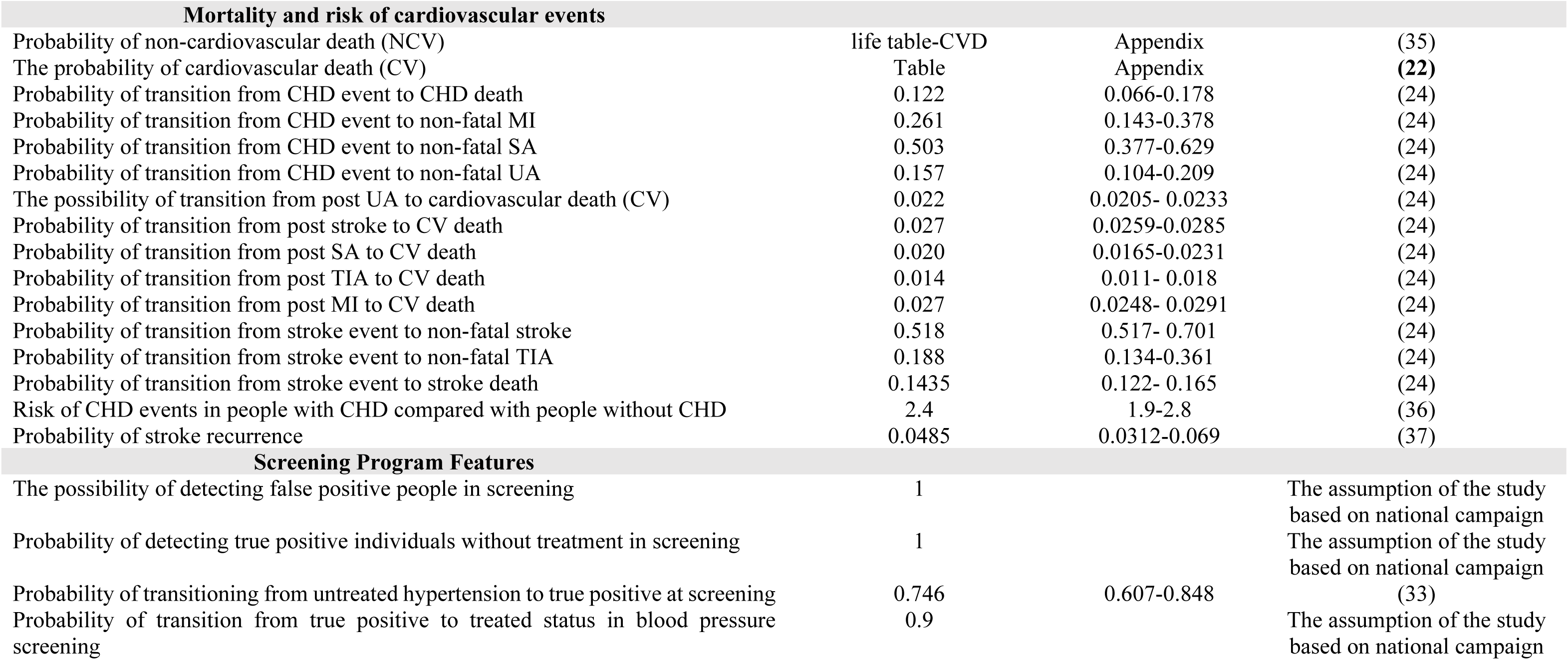
base-case model inputs.

For this purpose, the probability of disease incidence in the general population of the country was estimated by utilizing the relationship between the incidence rate and the probability of the disease (23). Subsequently, to estimate the probability of incidence in individuals with high blood pressure, the probability of incidence in the country’s population was multiplied by the risk of disease in individuals with high blood pressure, disaggregated by age groups.

Finally, the probability of incidence in these two groups, healthy individuals and those under treatment was also estimated by utilizing the risk of disease in each group. The risk of disease in different patient groups was extracted from prior studies(24).

Additionally, the probability of screening each individual within the target population during the national screening campaign was estimated using the results and data obtained from the national campaign.

#### Screening parameters

Data related to sensitivities, and specificity of screening tests were extracted and collected from prior studies. Additionally, some of the data including participant rate in the national campaign, were calculated based on the guidelines and results of the national hypertension control campaign (Table1).

#### Costs

**Costs of high blood pressure screening(For the general population),** These costs were calculated using data from the National High Blood Pressure Control Campaign, following a bottom-up approach. These costs were calculated at three levels, including healthcare centers, universities, and the Ministry of Health, and for estimation three groups of costs, including human resource costs, equipment costs, and consumable costs were considered;

**At the healthcare centers level,** human resources costs include the salaries and wages paid to employees and screeners present at blood pressure measurement stations. Based on the number of healthcare and non-healthcare (temporary) stations and the working hours at those stations, these costs were estimated.

In addition, equipment such as laptops, blood pressure monitors, tables and chairs, and examination beds were used to measure the blood pressure of pregnant women during the implementation of the screening program. The equipment cost includes the depreciation cost of the mentioned items during the screening program period.

Consumable costs included staff meals (lunch and dinner), snacks (fruit and biscuits), disposable cups, pamphlets, posters, banners, blood pressure registration cards, the clients’ information registration lists, and the cost of printing training certificates for screeners. The average cost of each item was multiplied by the required quantity for each station per day and then multiplied by the number of days in the program and the number of stations using it. After estimating and adding up the total cost of these items, the total cost of consumables was estimated.

**At the university level**, costs included meetings, visits, and coordination with relevant groups, which were collected through interviews with individuals involved at this level.

**At the Ministry of Health level**, costs included strategic committees, executive committees, and their working groups including communications and information dissemination, treatment group, training group, support group, social participation group, disease registration and identification group, and monitoring and evaluation group. The cost of these items was estimated using the National High Blood Pressure Control campaign guidelines and interviews with the executive committee (Deputy of Health, Ministry of Health).

#### Costs Of Diseases

In this study, data on the cost of high blood pressure and its related diseases were used. The costs of diagnosing and treating hypertension are explained in detail in the published study(25), and the cost of diagnostic tests for false positive individuals using treatment guidelines was calculated. For the costs related to heart disease and stroke, the prevalence and the number of cases of each disease using the IHME database for Global Burden of Disease were separately estimated(22), the cases of each disease were multiplied by the average cost per patient, and the total cost for each disease was calculated, considering new and previous cases, direct medical and non-medical expenses, as well as outpatient and inpatient costs.

Direct medical costs, encompassing high blood pressure diagnosis and treatment, along with costs related to selected diseases, Direct medical costs of selected diseases were calculated using health insurance data, survey(26) and a literature review(24, 27, 28), and Direct non-medical costs Using a literature review were calculated (27, 28). and indirect costs were calculated using the human capital approach Using the NASBOD and IHME database for Global Burden of Disease (22, 29). These costs were calculated using the bottom-up approach. then multiplied the costs for each disease category by the population-attributed fraction of high blood pressure and aggregated the costs related to high blood pressure. Data on demographic and economic data, such as population statistics, labor force employment rates, household activity rates, wage rates, and GDP per capita, were extracted from the Iran Statistics Center(30)and the World Bank(31). These expenses have been reported along with references in Table 2.

**Table 2:**
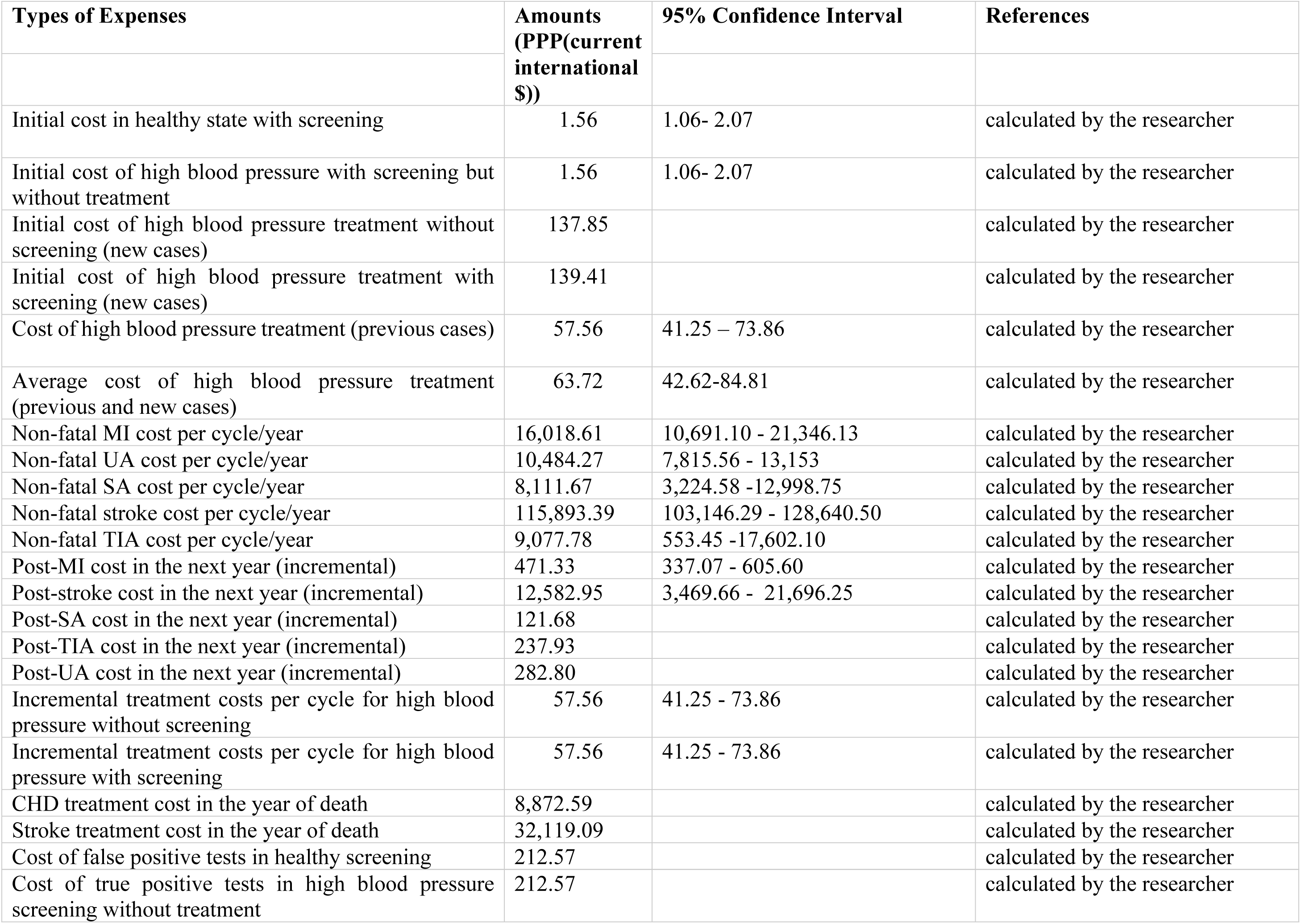
Estimated cost data for the model.

| **Types of Expenses** | **Amounts**<br>**(PPP(current** | **95% Confidence Interval** | **References** |
| --- | --- | --- | --- |
| | **international**<br>**\$))** | | |
| Initial cost in healthy state with screening | 1.56 | 1.06- 2.07 | calculated by the researcher |
| Initial cost of high blood pressure with screening but without treatment | 1.56 | 1.06- 2.07 | calculated by the researcher |
| Initial cost of high blood pressure treatment without screening (new cases) | 137.85 |  | calculated by the researcher |
| Initial cost of high blood pressure treatment with screening (new cases) | 139.41 |  | calculated by the researcher |
| Cost of high blood pressure treatment (previous cases) | 57.56 | 41.25 – 73.86 | calculated by the researcher |
| Average cost of high blood pressure treatment (previous and new cases) | 63.72 | 42.62-84.81 | calculated by the researcher |
| Non-fatal MI cost per cycle/year | 16,018.61 | 10,691.10 - 21,346.13 | calculated by the researcher |
| Non-fatal UA cost per cycle/year | 10,484.27 | 7,815.56 - 13,153 | calculated by the researcher |
| Non-fatal SA cost per cycle/year | 8,111.67 | 3,224.58 -12,998.75 | calculated by the researcher |
| Non-fatal stroke cost per cycle/year | 115,893.39 | 103,146.29 - 128,640.50 | calculated by the researcher |
| Non-fatal TIA cost per cycle/year | 9,077.78 | 553.45 -17,602.10 | calculated by the researcher |
| Post-MI cost in the next year (incremental) | 471.33 | 337.07 - 605.60 | calculated by the researcher |
| Post-stroke cost in the next year (incremental) | 12,582.95 | 3,469.66 - 21,696.25 | calculated by the researcher |
| Post-SA cost in the next year (incremental) | 121.68 |  | calculated by the researcher |
| Post-TIA cost in the next year (incremental) | 237.93 |  | calculated by the researcher |
| Post-UA cost in the next year (incremental) | 282.80 |  | calculated by the researcher |
| Incremental treatment costs per cycle for high blood pressure without screening | 57.56 | 41.25 - 73.86 | calculated by the researcher |
| Incremental treatment costs per cycle for high blood pressure with screening | 57.56 | 41.25 - 73.86 | calculated by the researcher |
| CHD treatment cost in the year of death | 8,872.59 |  | calculated by the researcher |
| Stroke treatment cost in the year of death | 32,119.09 |  | calculated by the researcher |
| Cost of false positive tests in healthy screening | 212.57 |  | calculated by the researcher |
| Cost of true positive tests in high blood pressure screening without treatment | 212.57 |  | calculated by the researcher |

In this study, all costs were adjusted to current international dollars based on the World Bank’s PPP conversion factors in 2020

(https://data.worldbank.org/indicator/PA.NUS.PPP?locations=IR).

#### Utility and Quality of Life Weights

Data related to utility or quality of life were extracted and collected from prior studies and is fully reported in Table 3.

**Table 3:**
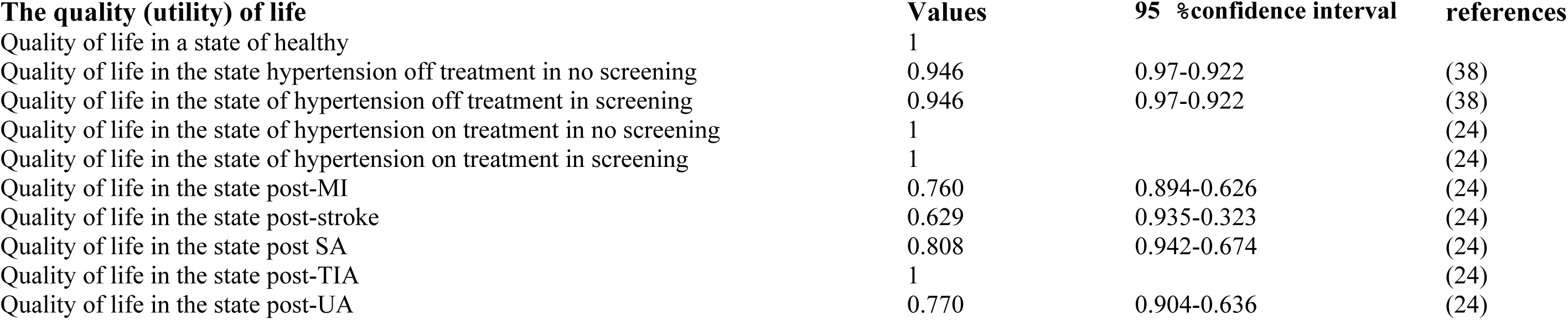
Data of utility and quality of life of the model.

| **The quality (utility) of life** | **Values** | **95 %confidence interval** | **references** |
| --- | --- | --- | --- |
| Quality of life in a state of healthy | 1 |  |  |
| Quality of life in the state hypertension off treatment in no screening | 0.946 | 0.97-0.922 | (38) |
| Quality of life in the state of hypertension off treatment in screening | 0.946 | 0.97-0.922 | (38) |
| Quality of life in the state of hypertension on treatment in no screening | 1 |  | (24) |
| Quality of life in the state of hypertension on treatment in screening | 1 |  | (24) |
| Quality of life in the state post-MI | 0.760 | 0.894-0.626 | (24) |
| Quality of life in the state post-stroke | 0.629 | 0.935-0.323 | (24) |
| Quality of life in the state post SA | 0.808 | 0.942-0.674 | (24) |
| Quality of life in the state post-TIA | 1 |  | (24) |
| Quality of life in the state post-UA | 0.770 | 0.904-0.636 | (24) |

After completing all the health states in the model, the model design process is nearly finished. Consequently, all initial health states and subsequent health states have been fully designed and model parameters were inserted. The cycles of the model were repeated for 70 periods (total life), each with a duration of one year in the Tree Age 2020 software. The simplified structure of the transition states in the Markov model is shown in Figure1(32).

In this model, after the screening, people are transferred from suspected to diagnosed states (people with normal blood pressure with a true negative or false positive test result, or high blood pressure with a true positive or false negative test result) and then people in this Health conditions remain or are transferred to a disease (event) during the time horizon (myocardial infarction, unstable angina, stable angina, stroke, transient ischemic attack and conditions after these diseases and death). In each case, the probability of each individual remaining in a particular state is determined using transition probabilities.

### Model implementation and Extraction of Findings

To implement the model, first the collected and calculated data were entered into the model.Then, the model’s output was evaluated for accuracy and conformity with reality using Markov Coherence in software, and any necessary changes were made to run the model. Afterward, the model was run to extract maps and cost-effectiveness ratios of screening strategies and other desired findings.

In this study, the Plane and cost-effectiveness ratio (incremental) of screening strategies using the Markov model in TreeAge software were extracted. By extracting the cost-effectiveness Plane, the cost-effectiveness frontier and dominated and non-dominated strategies were identified, as well as the willingness to pay chart and its intersection with the cost-effectiveness frontier. By extracting the incremental cost-effectiveness ratio of screening strategies, the status of each strategy was determined relative to no screening, relative to its previous best strategy, and relative to the cost-effectiveness threshold.

Also, the quantity of QALYs gained from identifying and preventing individuals from developing high blood pressure, coronary heart disease, ischemic stroke, hemorrhagic stroke, and their complications by implementing blood pressure screening strategies was estimated using the results of the Markov model. The value of each QALY was estimated based on the results of Jahanbin et al.’s study, which was 1.35 times the GDP per capita(39). This amount is less than the estimated threshold for cancer interventions(40). Additionally, the GDP per capita was extracted from World Bank reports(41).

The benefits of these strategies include the monetary value of saved life-years (the number of QALYs gained), and to calculate the net benefit, we subtracted the costs of each strategy from these benefits. The costs of the program include the intervention costs and the costs of the disease and its complications.

### Sensitivity analysis

In this study, various sensitivity analyses were conducted to test the strength of the assumptions of the model and data sources. First, a one-way sensitivity analysis was performed between important parameters that had the most significant impact on the results in each screening strategy.

The variables included in this analysis were the “Probability of transition from coronary heart disease to non-fatal stable angina, quality of life after stable angina, probability of transition from coronary heart disease to non-fatal unstable angina, quality of life after unstable angina, quality of life after myocardial infarction (MI), the discount rate of outcomes, cost of stroke in the next year, quality of life after stroke, the discount rate of costs, probability of transition from healthy to true negative in screening, probability of transition from stroke to non-fatal transient ischemic attack, probability of transition from high blood pressure without Treatment to true positive in screening, cost of nonfatal stroke, probability of stroke recurrence and coronary heart disease, probability of transition from stroke event to nonfatal TIA, and probability of transition from CHD to CHD-related death”. The sensitivity analysis involved three scenarios: one with base values, another with minimum possible values for parameters, and a third with maximum possible values. This approach allowed for an evaluation of how results responded to variations at both ends of these key variables, providing insights into the model’s sensitivity across different scenarios.

Furthermore, a probabilistic sensitivity analysis was used to quantify the uncertainty around the point estimates of input parameters. A probability distribution was defined for each input parameter of the model. The model was then run multiple times (1000 iterations), each time a value for each input parameter was randomly sampled from its corresponding probability distribution. The average costs and QALYs were calculated using these sampled values. The probability distribution in the analysis was based on error estimates from prior studies. The results of the probabilistic sensitivity analysis (PSA) were presented in cost-effectiveness acceptability curves, scatter plots, and cost-effectiveness acceptability curves for willingness-to-pay thresholds.

## Results

As indicated in Figure 1, all screening strategies have higher costs and effectiveness compared to no screening. Non-dominant strategies are connected to each other by a line that forms the cost-effectiveness frontier.

**Figure 1:**
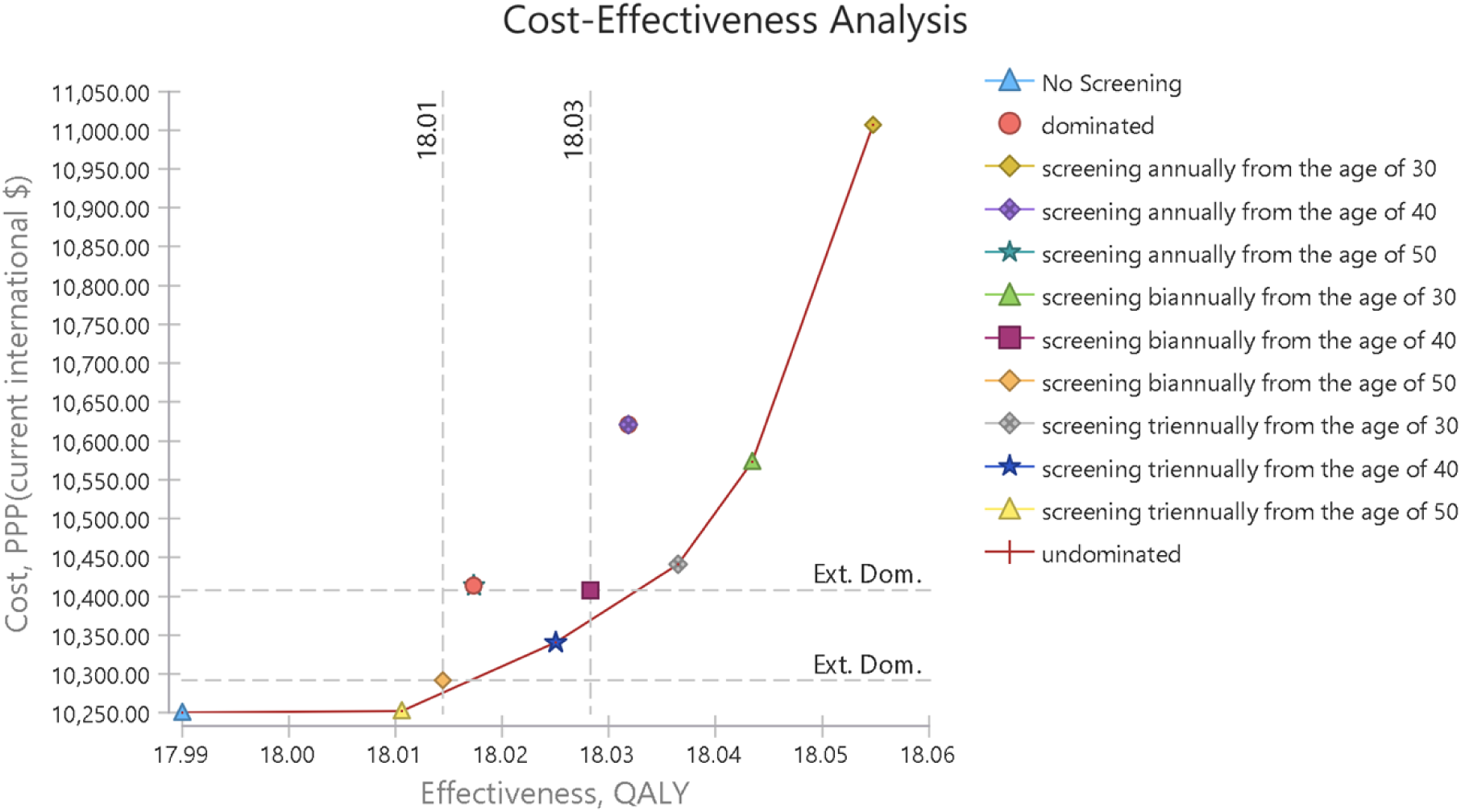
Cost-Effectiveness Plane of Screening Strategies.

The cost-effectiveness frontier and out-of-frontier strategies are evident in Figure 1, and as shown in this graph, among the 9 different strategies and non-screening, five interventions are undominated(non-dominated), while the annual screening strategies starting at age 40 and age 50 are absolutely dominated, and the biennial screening strategies at both age 40 and 50 are Extended dominated.

In addition, in Figure 2, the cost-effectiveness plane is shown along with the willingness-to-pay curve. The willingness-to-pay curve shows the slope of the willingness to pay, which is tangent to the optimal strategy. The tangency of the willingness-to-pay slope with any strategy indicates the optimality or the high probability of cost-effectiveness of that intervention, and it helps in selecting a cost-effective strategy. Of course, only the cost-effective frontier strategies can be optimal choices. In this graph, the willingness-to-pay slope is tangent with the strategy of screening every two years from the age of 30. In Table 4, the costs and incremental benefits of all screening strategies compared to no screening have been reported. According to this table, all screening strategies have higher costs and greater effectiveness compared to no screening.

**Figure 2:**
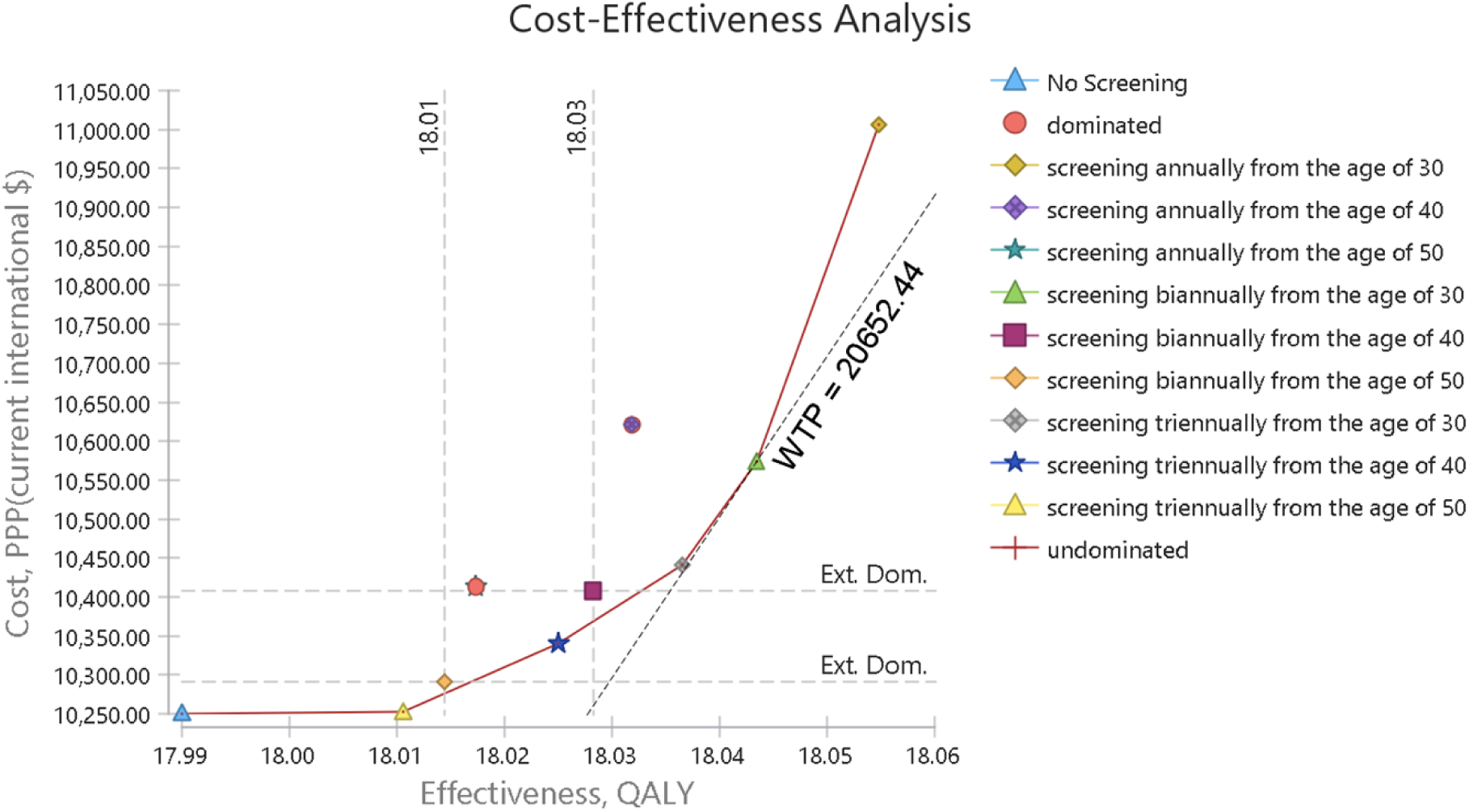
Cost-effectiveness plane and willingness to pay.

**Table 4:**
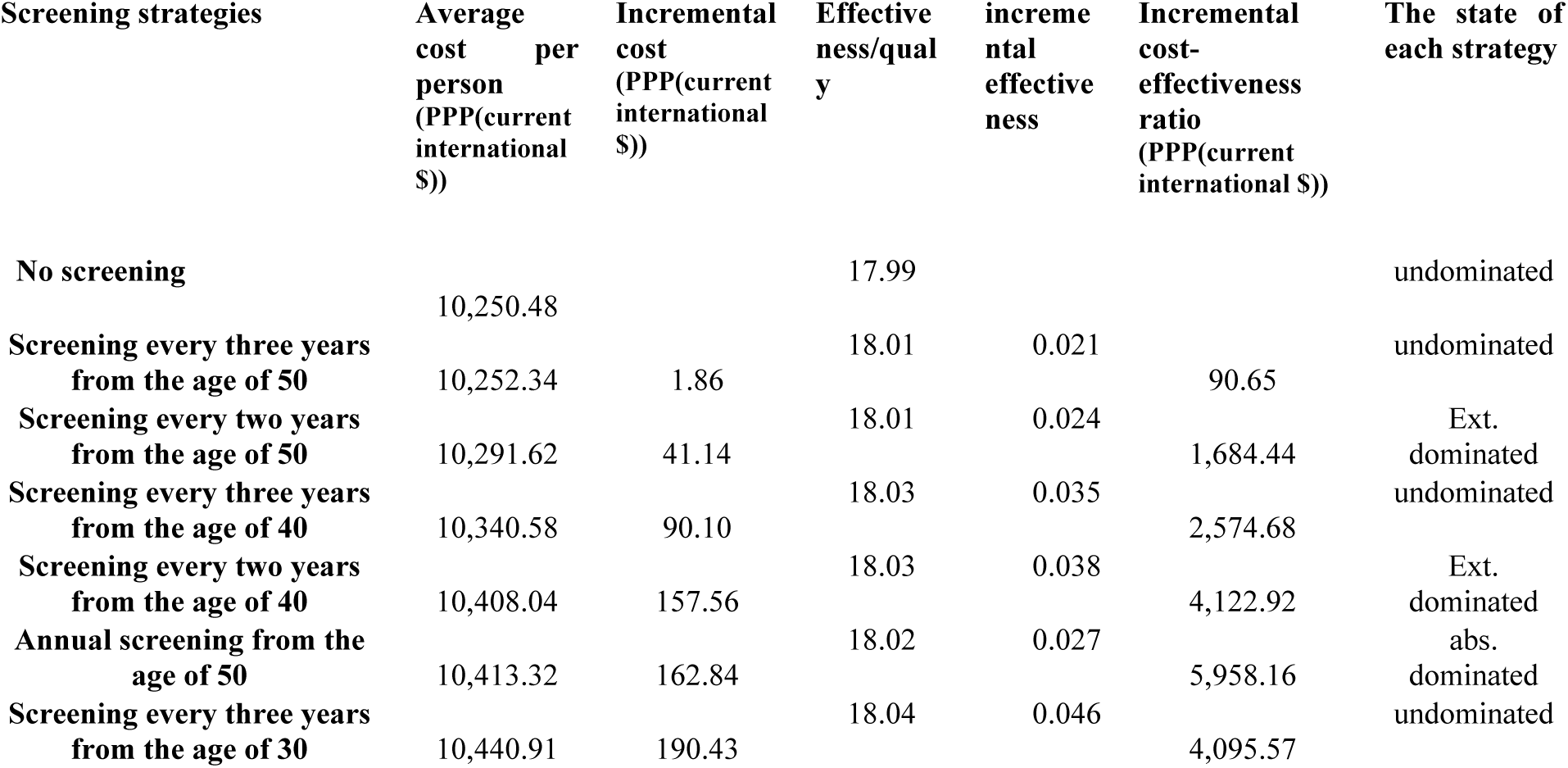
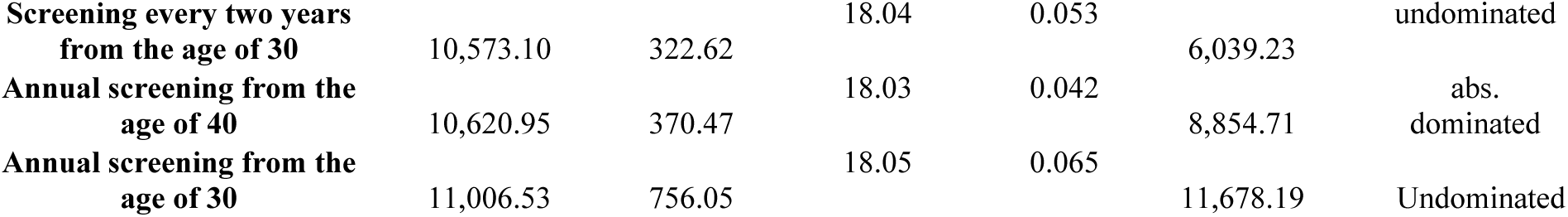
Incremental costs and benefits of screening strategies compared to no screening.

Additionally, in Table 5, based on the results of the Markov model, the incremental cost-effectiveness ratios (ICERs) of undominated screening strategies compared to their previous best strategy have been compared and ranked. In this table, four dominated strategies have been eliminated, and only the undominant strategies have been ranked in order of increasing effectiveness. After eliminating the dominated strategies, five strategies remained, including screening every three years from ages 50, 40, and 30, and screening every two years and annually from age 30. The ICERs of these strategies, compared to their own previous best strategies, were $ PPP 90.65, 6,114.45, 8,723.89, 19,089.06, and 38,289.63, respectively.

**Table 5:**
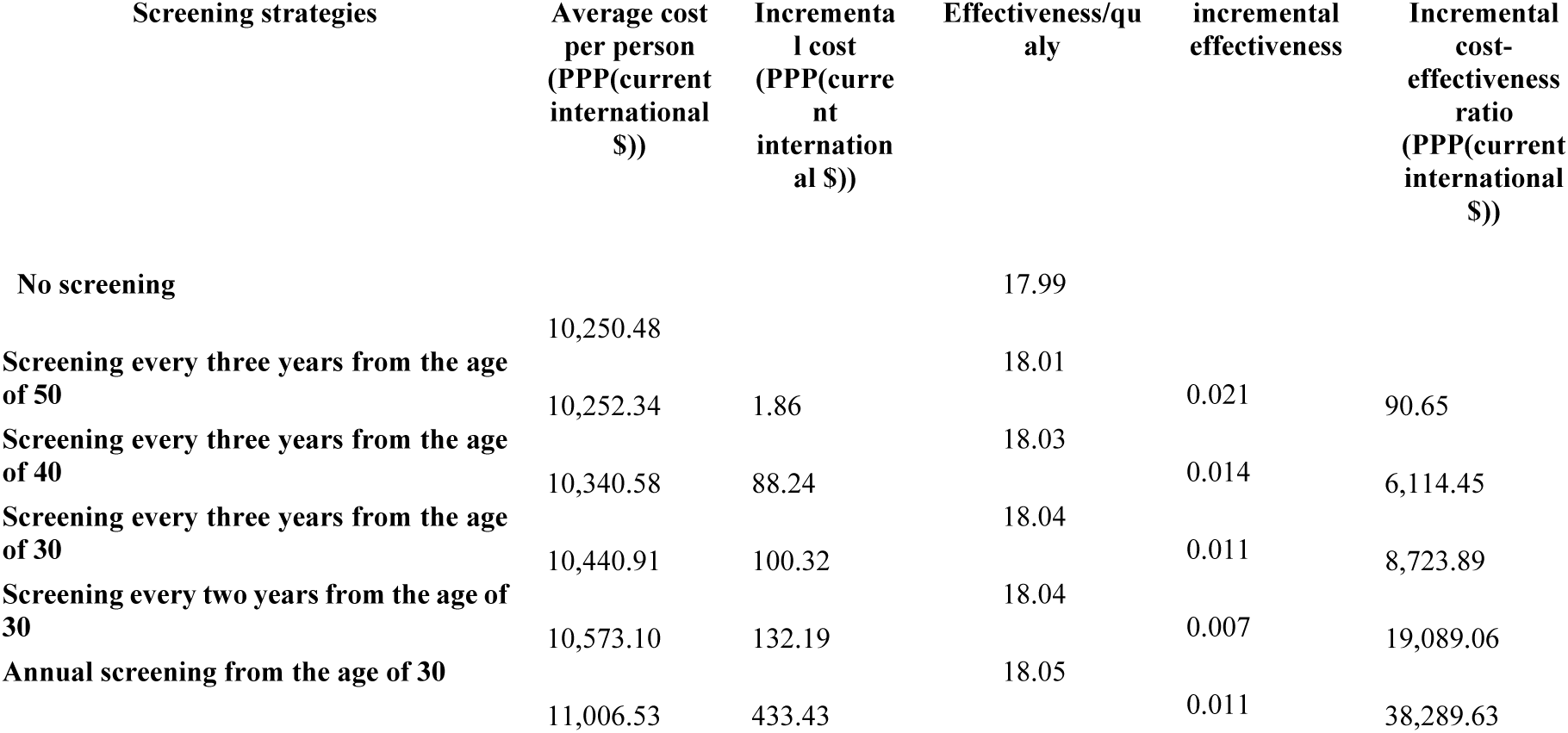
Costs and benefits of non-dominated screening strategies.

Furthermore, by implementing screening interventions, in all strategies, a certain amount of gained quality-adjusted life years (QALYs) has been added to each individual throughout their remaining lifetime compared to no screening. In Table 4, the number of gained QALYs per individual for each blood pressure screening strategy compared to no screening has been reported, and in Table 5, the gained QALYs compared to the previous best strategy have been reported for non-dominant cases.

### sensitivity analysis

We performed various sensitivity analyses to test the robustness of the model results. First, we conducted a univariate sensitivity analysis between key model parameters (including discount rates for costs and outcomes, some costs, quality of life, and probabilities), and the results of this sensitivity analysis were presented using tornado diagrams. The title, baseline values, and the range of selected variables for the sensitivity analysis of different screening strategies are presented in the following table.

**Table 8:** Selected variables for one-way sensitivity analysis.

| Variables | Amounts (costs in PPP(current international \$)) |
| --- | --- |
| Cost of non-fatal stroke | 115,893.39 |
|  | (95%CI:103,146 -128,641 |
| Cost of post stroke in following year | 12,582.95 |
| Discount rate for costs | 0.05 (0.03-0.07) |
| Discount rate for outcomes | 0.05(0.03-0.07) |
| Probability of Stroke Recurrence | 0.05(0.031-0.069) |
| Quality of life in the state post MI | 0.76(95%CI:626-894) |
| Quality of life in the state post stroke | 0.63(95%CI:323-935) |
| Quality of life in the state post SA | 0.81(95%CI:674-942) |
| Quality of life in the state post UA | 0.77(95%CI:634-904) |
| Quality of life in the state hypertension off treatment | 0.946(95%CI:0.922-0.97) |
| relative risk for CHD events in those with established CHD compared with CHD-free | 2.40(1.9-2.8) |
| Transition probability from chd event to non fatal SA | 0.50(0.377-0.629) |
| Transition probability from chd event to non fatal UA | 0.16(0.104-0.209) |
| Transition probability from healthy to true negative in screening | 0.75(95%CI:0.479-0.904) |
| Transition probability from hypertension off treatment to true positive in screening | 0.75 (95%CI:0.607-0.848) |
| Transition probability from Stroke event to non fatal TIA | 0.19(0.134-0.361) |
| Transition probability from chd event to CHD death | 0.122(0.066-0.178) |

According to the deterministic sensitivity analysis results, in most strategies, the estimated outcomes were more sensitive to changes in several variables, including “probability of transition from healthy to true negative individuals (Specificity of screening test), discount rates for outcomes and costs.” In this model, probabilistic sensitivity analysis (PSA) was also used to account for uncertainty around the point estimates of input parameters. The PSA results showed very small differences between the estimated average costs and qalys (effectiveness) and their values in the base case analysis, indicating the stability of the results. The PSA results were presented using cost-effectiveness acceptability curves, acceptability at the willingness-to-pay Curve, and cost-effectiveness scatter plots.

The cost-effectiveness acceptability curve is a commonly used output for communicating the results of probabilistic sensitivity analysis in cost-effectiveness models. The acceptability curve shows the relative cost-effectiveness as a function of the willingness-to-pay threshold. For each willingness-to-pay value, the curve uses net benefits to determine the percentage of simulation iterations that are deemed appropriate for each strategy. As the willingness-to-pay threshold increases, the percentage of iterations for more effective strategies increases (Briefly, this curve indicates the probability of cost-effectiveness at any point within a range or domain of willingness-to-pay). Like a sensitivity analysis, the acceptability curve requires a wide range of values for willingness-to-pay (ICER threshold).

Furthermore, Figure 3 displays the cost-effectiveness acceptability curve that shows the probability of cost-effectiveness at different willingness-to-pay thresholds. In other words, this graph shows the probability of each strategy being cost-effective at the specified threshold.

**Figure 2:**
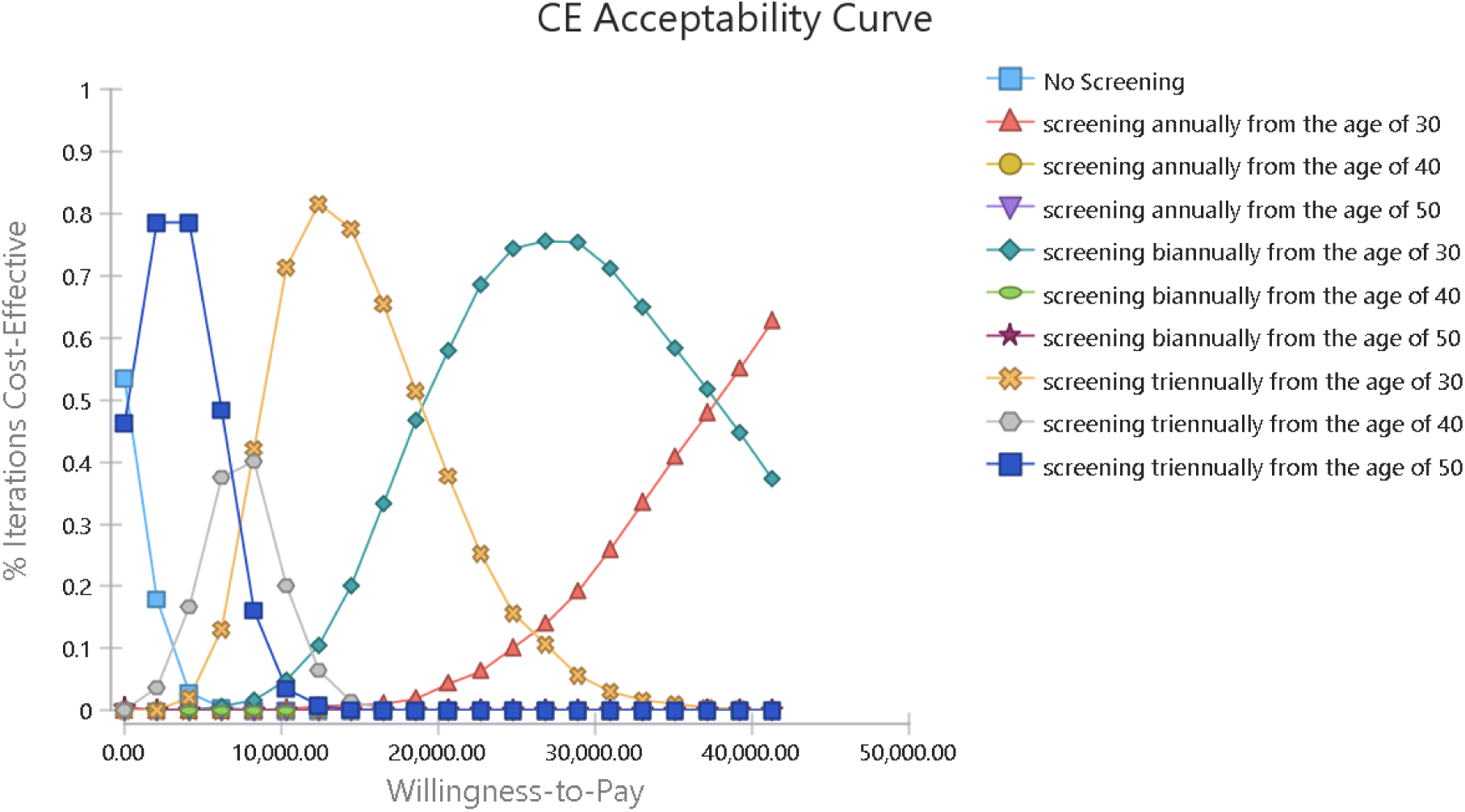
Cost-Effectiveness Acceptance Curve.

**Figure 3:**
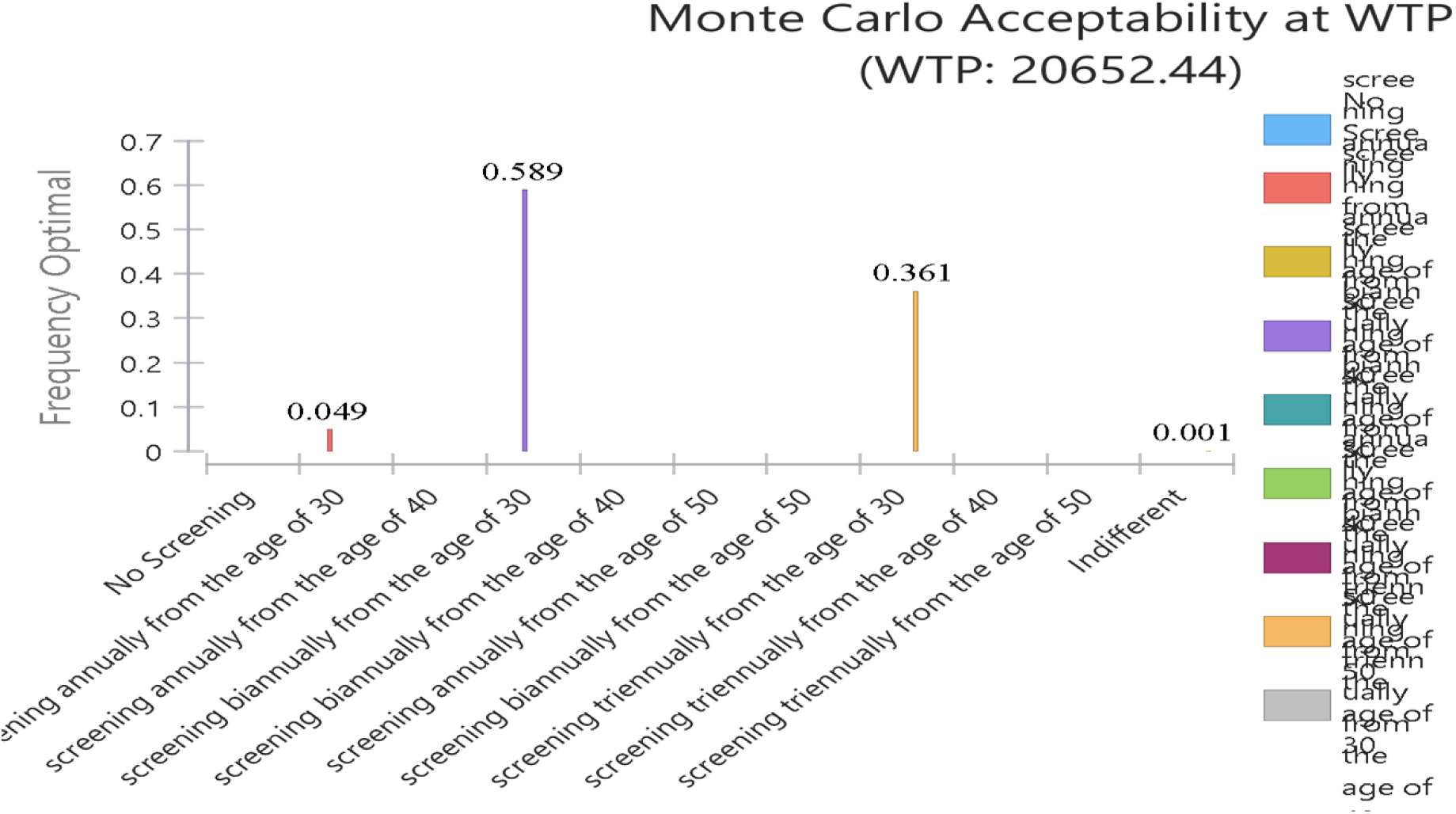
Acceptability at Willingness-to-Pay Threshold in Probabilistic Sensitivity Analysis.

According to Figure 3, at a threshold of 1.35 times GDP per capita ($ PPP 20,652.44), the probability of cost-effectiveness for screening every two and three years from age 30 was 0.589 and 0.361, respectively, and the probability of cost-effectiveness for annual blood pressure screening from age 30 was 0.049, while the probability of cost-effectiveness for other strategies was almost zero.

**Figure 4:**
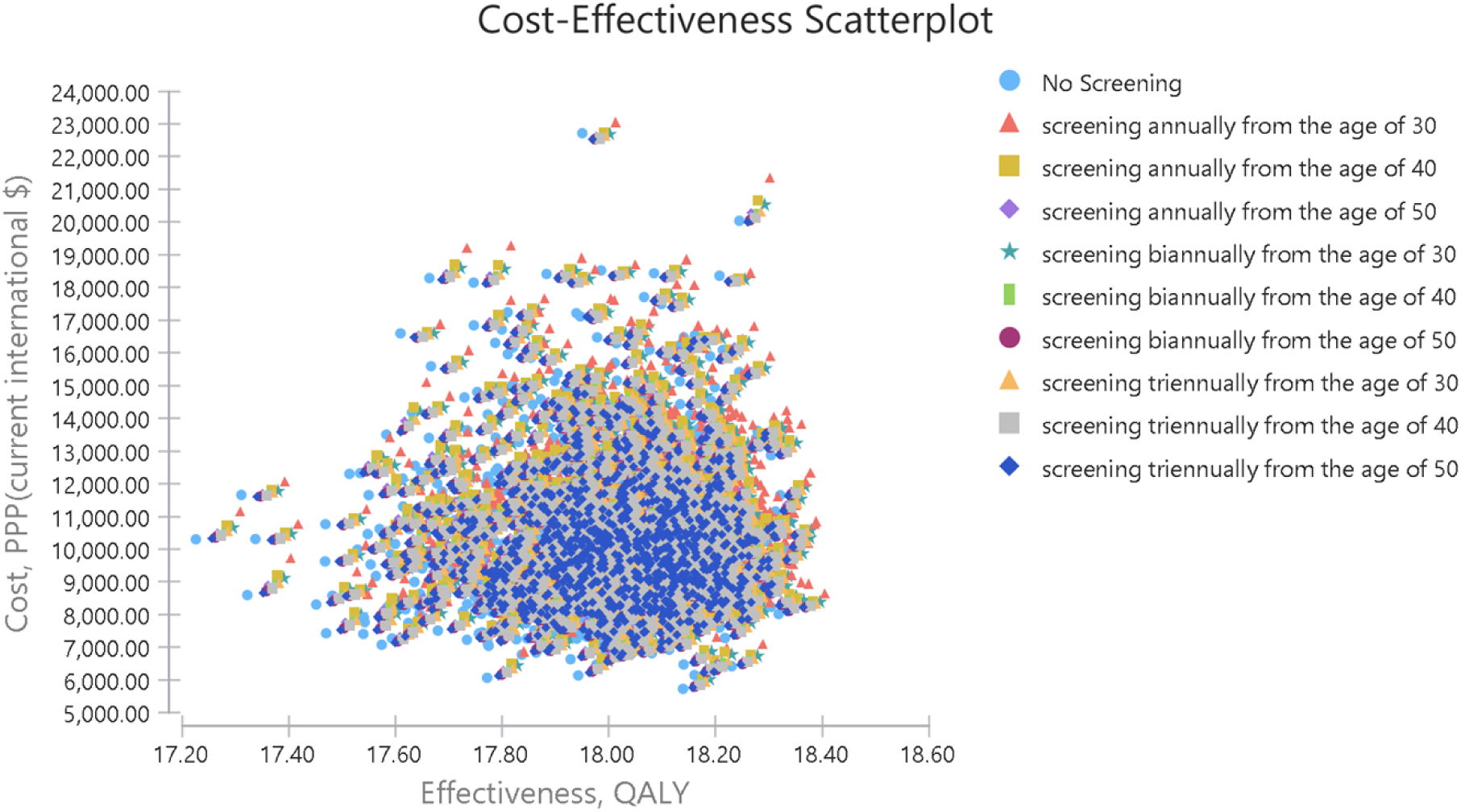
Cost-Effectiveness Scatter Plot.

The cost-effectiveness plane, which plots each strategy’s average cost and effectiveness, can naturally expand to a scatter plot for simulation. The scatter plot uses the cost-effectiveness plane to plot pairs of costs and effects separately for each model recalculation. all points of each strategy have a specific color.

## Discussion

In this study we analyzed the economic evaluation of nine blood pressure screening strategies, and the cost-effectiveness ratios and other results of the screening strategies were reported in two groups. In one group, an incremental cost-effectiveness ratio was reported for the screening strategies compared to no screening, and all screening strategies had an incremental cost-effectiveness ratio less than the threshold or willingness-to-pay (($ PPP 20,652.44). However, four strategies, including annual screening from age 40 and 50 due to higher costs and lower effectiveness (absolutely dominated), and screening every two years from age 40 and 50 because of a higher incremental cost-effectiveness ratio compared to other interventions, were dominated.

The second group comprises five strategies, excluding dominated cases. Four of these involve screening every three years at ages 40, 30, and 50, and every two years from age 30, with an incremental cost-effectiveness ratio ranging from $PPP 90.54 to $PPP19,089.02 per Quality Adjusted Life Year (QALY) gained, which is less than the threshold cost-effectiveness ratio (($ PPP 20,652.44). And the last strategy is annual screening from age 30, which had an incremental cost-effectiveness ratio greater than the threshold.

According to a study conducted in Korea in 2021, the cost per Quality Adjusted Life Year (QALY) with hypertension screening strategies was less than the incremental cost-effectiveness ratio threshold (approximately 30.5 million KRW) compared to no screening. Strategies, including the first screening with confirmatory examination every three years for adults over 40 (10.2 million KRW), every two years (13.2 million KRW), or annually (19.9 million KRW), were deemed cost-effective. The most cost-effective strategy involved a first screening with a second confirmatory examination at age 40 or older every three years(15).

Additionally, in the study conducted in Vietnam, screening scenarios varied in intervals (one-time, annually, biennially) and initiation ages (35, 45, or 55 years), considering treatment coverage. Over a 10-year and lifetime horizon, probabilistic sensitivity analysis addressed parameter uncertainty, with a decision-making threshold set at three times the per capita gross domestic product (GDP).

According to the screening results, for men starting screening at the age of 55, all screening scenarios were likely to be cost-effective. For women starting screening at the age of 55, the one-time screening had a 90% chance of being cost-effective. Over a lifetime horizon, the cost per quality-adjusted life year (QALY) obtained was less than the threshold of 15,883 international dollars in all screening scenarios among men. Similar results were observed for women starting screening at the age of 55. Additionally, if biennial screening with an increase in treatment coverage to 20% is considered or even with only biennial screening, starting screening in women at the age of 45 is likely to be cost-effective(19).

Routine blood pressure screening in adolescents is effective, but population-based interventions with broader accessibility for preventing cardiovascular diseases may offer greater cost-effectiveness and efficiency. It is recommended to select and implement interventions based on their cost-effectiveness ratio, considering community conditions and feasibility(42).

Various studies have demonstrated the cost-effectiveness of interventions related to screening, control, and management of high blood pressure, education, and self-monitoring (15, 36, 37). The cost-effectiveness of blood pressure screening has been emphasized in studies conducted on national and routine screening programs as well as population-based screening(14). Screening potentially reduces diagnosis and treatment time and can be cost-effective if it is linked to primary healthcare through health centers for providing treatment to patients identified through screening (17).

Integrating high blood pressure screening into routine medical examinations and health insurance coverage is deemed appropriate from a health economics perspective.Preventing cardiovascular diseases through high blood pressure screening is cost-effective. Screening strategy should be based on age, gender, and screening intervals(18). A comprehensive approach to care and prevention, facilitated by government support, subsidies, and an active health network operating at different service levels, has the potential to be cost-effective in a screening program(38).

In the present study, the net present value and incremental net present value of all blood pressure screening interventions for each individual with a 5% discount rate were positive. The incremental net benefits of screening every two and three years from age 30 was significantly higher than the other strategies, amounting to $PPP780.65 and $PPP769.82, respectively, while in the strategy of annual screening from the age of 30, it was $PPP581.

The study also revealed positive base and annual return on investment (ROI) for all screening interventions. The base ROI was higher due to not considering the compound effect, which could create a significant difference over time. The longer the time period, the greater the difference between the approximate annual ROI (base), which is calculated by dividing ROI by the study period, and the annualized return on investment.

A study in Ghana assessed the cost-benefit of hypertension screening and treatment by community health workers. The analysis, based on screening 25,000 individuals above the age of 30, revealed that with a diuretic treatment regimen and a 30% long-term adherence rate, the intervention could prevent 29 deaths (equivalent to 505 years of life), 11.1 cases of heart disease, 0.1 cases of stroke, and 1.6 cases of heart failure over ten years. The estimated benefits amounted to 7.1 million ¢GH (1.6 million dollars) at an 8% discount rate, with intervention costs of 2.2 million ¢GH (0.5 million dollars), resulting in a benefit-to-cost ratio of 3.3(43). These findings align with the present study’s results, suggesting that screening interventions can be implemented with higher effectiveness and efficiency, or as part of multi-interventional preventive programs for managing and treating high blood pressure(40) (41). Considering existing healthcare networks and infrastructure, such interventions not only identify undetected individuals through health systems but also raise awareness about the disease, especially among those at risk. With a cohesive healthcare referral system, diagnosing and treating individuals becomes more feasible, making preventive interventions like blood pressure screening highly effective.

In the annual screening strategy for individuals over 30, the average cost per person screened, diagnosed with high blood pressure, or suspected of having high blood pressure, was $PPP1.56, $PPP5.56, and $PPP46.80, respectively. Treating each diagnosed patient incurred a cost of $PPP137.85, while the net benefit of screening strategies compared to no screening ranged from approximately $PPP581 to $PPP780.65. The budgetary impact analysis projected a total cost of $PPP 438,799,413.74 for high blood pressure screening, diagnosis, and treatment interventions for individuals over 30 in the first year.

Considering the budgetary impact of screening interventions compared to the consequences resulting from untreated hypertension, prioritizing screening and treatment interventions for patients with high blood pressure under the leadership of the Ministry of Health could yield significant benefits. The study’s results are based on the economic evaluation of high blood pressure screening, and it suggests that programs conducted with broad participation from national institutions and media, particularly radio and television, not only directly benefit patients by identifying them through screening but also create awareness and sensitivity among the public, encouraging active participation in diagnostic and treatment processes. These additional benefits were not evaluated in the study.

The study demonstrates that screening and treatment strategies for hypertension, when implemented within a comprehensive and integrated framework, can be cost-effective, especially at a willingness-to-pay threshold of 1.35 times the GDP per capita per Quality-Adjusted Life Year (QALY), equivalent to $PPP 20,652.44 per QALY. This approach yields significant benefits for both the population and the government. Probabilistic sensitivity analysis indicates that biennial and triennial screening from age 30 have the highest probabilities of cost-effectiveness at this threshold, with probabilities of 0.589 and 0.361, respectively. Annual screening from age 30 also has increasing cost-effectiveness probabilities as the willingness-to-pay threshold rises.

In the present study, probabilistic sensitivity analysis demonstrates very low uncertainty in the base case analysis, indicating that the study findings are robust to a large extent.

## Limitations

The screening costs provided in the study lack some necessary information such as the expenses associated with building or renting healthcare centers and the cost of some services. Furthermore, some costs were derived from previous studies, which may differ from current patient costs.

The Markov model was designed based on the assumption that the follow-up and treatment of suspected and diagnosed individuals align with the screening program guidelines. However, in reality, not all eligible individuals may receive complete follow-up and treatment. While the study acknowledges this limitation and adjusts the model accordingly, its conclusions still rely on adherence to screening guidelines.

## Conclusion

High blood pressure is one of the most dangerous and very costly chronic diseases, but it is preventable. Preventive interventions like blood pressure screening can be the optimal and cost-effective strategies for controlling its economic burden. The study results identify screening every two and three years from the age of 30 as the optimal strategy, with probabilities of cost-effectiveness at 0.589 and 0.361, respectively.

Early and universal measures are deemed necessary to swiftly prevent and treat high blood pressure. Failure to address this health issue may lead to increased mortality, reduced life expectancy, and a substantial economic burden on both individuals and the government.

Policymakers and health system planners are urged to create favorable conditions for various preventive interventions. This includes initiatives such as education and awareness campaigns to encourage regular blood pressure measurement and maintaining blood pressure at optimal levels. Implementing these measures is essential for reducing the prevalence of high blood pressure and improving overall public health outcomes.

## Data availability statement

All data can be provided based on a reasonable request and with the approval of the Ethics Committee of Tehran University of Medical Sciences.

## Patient consent for publication

Not applicable.

## Ethics approval

This study is a part of the doctoral dissertation and has been approved by Tehran University of Medical Sciences ethics committee (IR.TUMS.VCR.REC.1399.380).

## Competing Interests

The authors declare no support from any organization for the submitted work.

## Acknowledgements

This study is a part of the doctoral dissertation of health economics. We hereby thank and appreciate the support of the Tehran University of Medical Sciences for its approval and support.

## Contributors

RD, was the thesis adviser and had the main role in modeling, data analysis, and making corrections in writing the paper. AA, the supervisor of the thesis, helped in analyzing the data and checking the correctness of the study. MZ, collection, modeling, analysis of data, and writing the article. EY, helping in some data collection.

## Funding

This study is a part of the doctoral dissertation and was done with the support of Tehran University of Medical Sciences.

## Notes

### Competing Interest Statement

The authors have declared no competing interest.

### Funding Statement

The author(s) received no specific funding for this work.

